# Two New Improved Methods for Determination of Cobb’s Angle of Scoliosis

**DOI:** 10.1101/2020.06.11.20124735

**Authors:** Dietrich W. Hellen, Michael Teusner

## Abstract

Cobb’s traditional method for measuring the angle of scoliosis is complicated and error-prone in application. Angles measured with this method are inaccurate and it therefore stands to reason to search for improved procedures.

We propose Cobb’s angle to be determined by two novel methods: the *distance method* and the *two-angle method*. Using mathematical error analysis and experimental test measurings we demonstrate that the two proposed methods offer significant advantages over the current standard. They simplify the measurement process and increase outcome accuracy.

For Cobb’s angles larger than 30 degrees, the two-angle method reduces the error-variation by approx. 25%. For Cobb’s angles below 30 degrees the distance method reduces the error-variation by approx. 50%.

## 1. Introduction

Cobb’s angle [1] is the fundamental measuring parameter for determining the degree of curvature in scoliosis. For determination of Cobb’s angle, two straight tangent lines along the end plates of the end vertebrae of the curvature are needed. Generally, Cobb’s angle α is measured at the point of intersection between two perpendicular lines drawn at these tangent lines (see Figure 1). However, application of this traditional procedure results in a considerable variation of error [2 − 10]. For small angles (mostly associated with static scoliosis) this variation of error can even reach the size of the angle itself. Thus, the measured angles can be totally wrong and the need for improved procedures is obvious.

**Fig 1.**
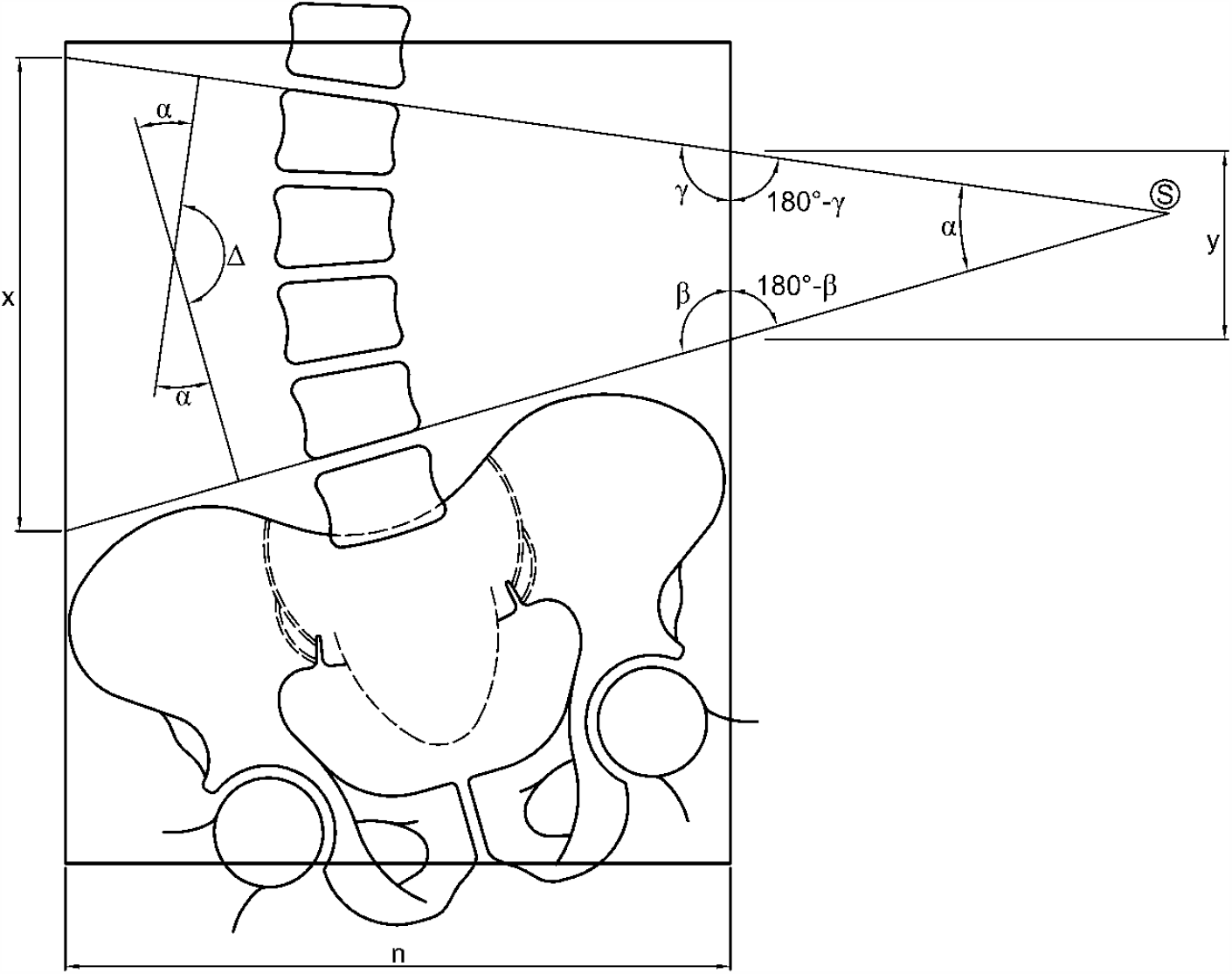
Measuring data for the different calculation methods.

## 2. Methods

### 2.1 Description of the Methods

In all imaging methods for the determination of Cobb’s angle, first the two tangent lines need to be established at the end plates of the inferior and superior end vertebrae (see Figure 1).

Therefore, the first two steps can be described for all methods as follows:

Step 1: Drawing the tangent line to the superior end plate of the superior end vertebra.

Step 2: Drawing the tangent line to the inferior end plate of the inferior end vertebra.

The various procedures differ in the following steps 3 to 5. These differing steps are described below (see Figure 1).

#### 2.1.1 Standard Cobb Method (Four-Line Method)

Step 3: Drawing a perpendicular line to the superior tangent line on the radiograph next to the image of the spine.

Step 4: Drawing a perpendicular line to the inferior tangent line on the radiograph next to the image of the spine in such a way that it intersects with the upper perpendicular line.

Step 5: Reading Cobb’s angle *α* from a protractor at the point of intersection of the two perpendicular lines.

#### 2.1.2 Two-Angle Method

Step 3: Measuring the angle *β* between the inferior tangent line and the side edge of the radiograph.

Step 4: Measuring the angle *γ* between the superior tangent line and the side edge of the radiograph.

Step 5: Computing of Cobb’s angle *α* by using:

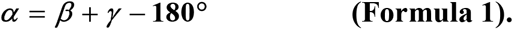

#### 2.1.3 Distance Method

Step 3: Measuring (with a ruler) the distances *x* and *y* between the points of intersection of the tangent lines with the side edges of the radiograph.

Step 4: Computing the difference *d* = *x* − *y*.

Step 5: Computing Cobb’s angle *α* by using:

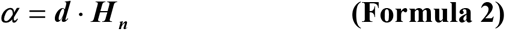

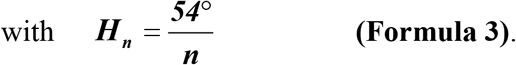

### 2.2. Explanation of the Methods

#### 2.2.1 Standard Cobb Method (Four-Line Method)

For the angle *α*_*s*_ at the point S of intersection in Figure 1 we have:

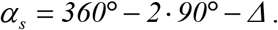

At the same time, the angle Δ at the point of intersection of the lines perpendicular to the tangent lines satisfies the equation:

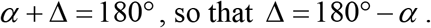

By introducing this term into the equation for *α*_*s*_ we obtain

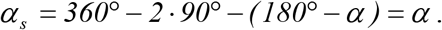

Hence the angle *α* at the point of intersection of the lines perpendicular to the tangent lines is identical to Cobb’s angle *α* _*s*_ at the point S of intersection of the tangent lines.

#### 2.2.2 Two-Angle Method

Cobb’s angle *α* can be determined without the construction of perpendicular lines to the tangent lines (as necessary in the Cobb Method). For this purpose one must determine the angles *β* and *γ* at which the tangent lines meet the side edge of the radiograph. With these angles, the sum of the angles in the triangle outside the radiograph (see Figure 1) satisfies the following equation:

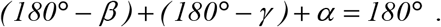

Thus we have:

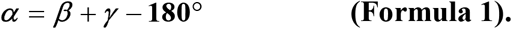

This equation is valid for all film formats and all angles of curvature of scoliosis, if the tangent lines intersection point Slies outside of the radiograph. Should the tangent lines intersect on the radiograph, Cobb’s angle *α* can be read directly at point S.

#### 2.2.3 Distance Method

The Distance Method allows determination of Cobb’s angle without using a protractor. For the description of this simple procedure we refer to Figure 2. Cobb’s angle *α* is composed of the two partial angles *α*_1_ and *α*_2_. By parallel shifting the line segment BC into the line segment AD, the angle α is moved (as a congruent step angle) from point S to point A. The two partial angles *α*_1_ and *α*_2_ are now located in rectangular triangles, namely *α*_1_ in EAF and *α*_2_ in EAD. The line segment EA equals the width of the radiograph *n* so that:

**Fig 2.**
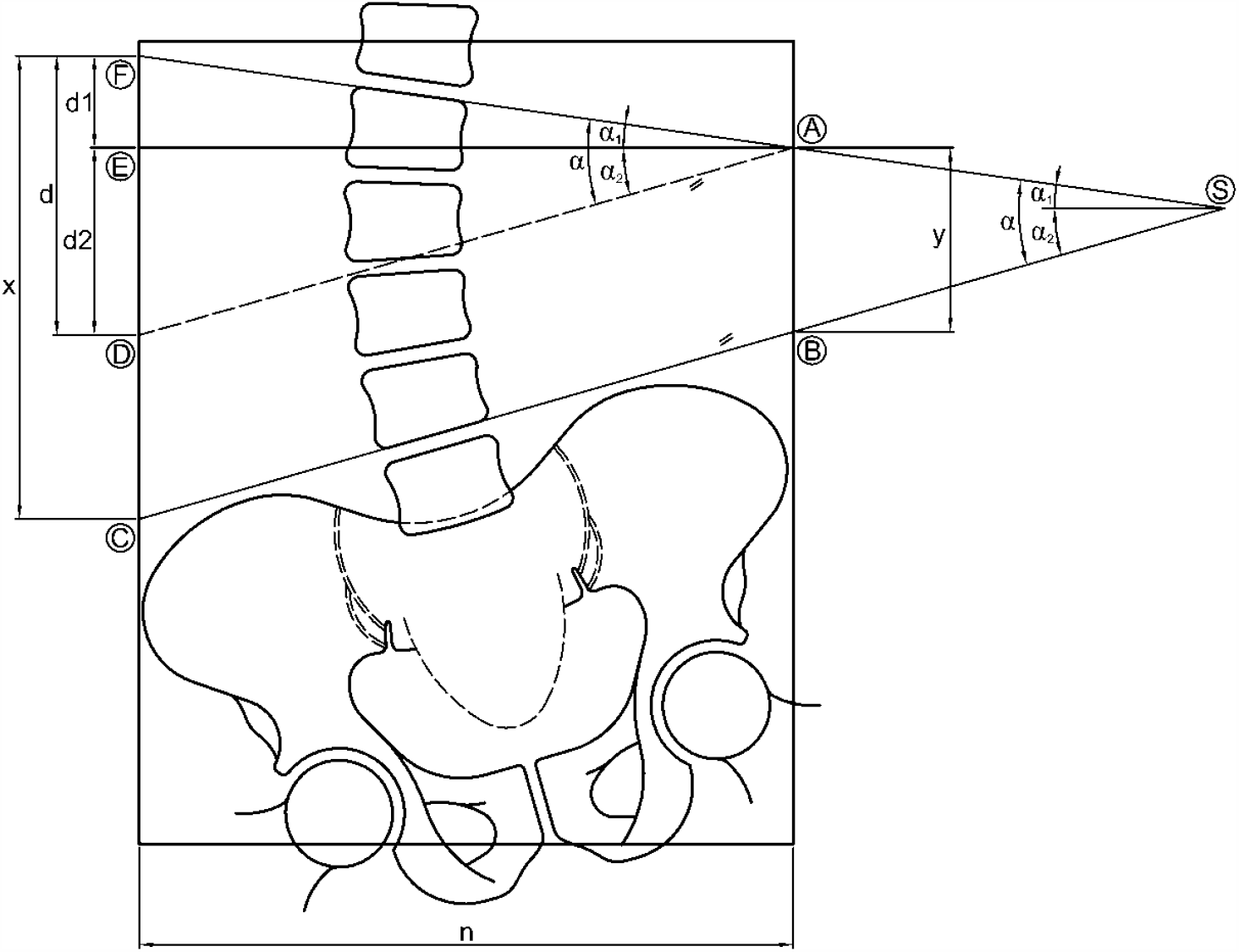
Explanation of the Distance Method.

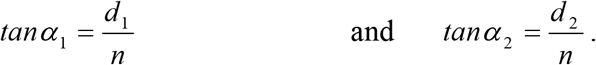

Hence:

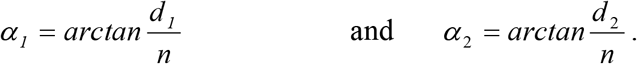

As *α* = *α*_1_ *+ α*_*2*_ we have:

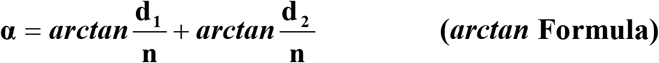

As the accurate computation of *α* by the *arctan* Formula would be complex, it will not be used for the Distance Method directly. Instead of this, a simplified formula shall be applied. For small angles (up to 30°) the *arctan* function is almost linear. Due to this fact, as long as the fractions 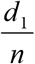 and 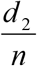 in the *arctan* Formula are sufficiently small, the *arctan* function can be replaced by an appropriate linear function as follows:

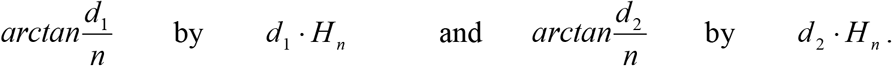

Here *H*_*n*_ is a constant corresponding to the film width *n*. Linearization simplifies the *arctan* Formula for *α* so that

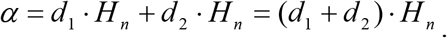

The sum of the distances *d*_1_ and *d*_2_ equals the distance *d*, which in turn equals the difference between *x* and *y*. Hence

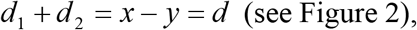

with *d* being the difference of the distances between the points of intersection of the tangent lines at the right and left side edges of the radiograph, respectively.

Thus,

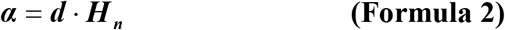

For any film width *n* the film’s characteristic parameter *H*_*n*_ is defined in such a way that the tolerable error occurring in linearizing the *arctan* function is as small as possible. This is the case, if *H*_*n*_ is calculated according to

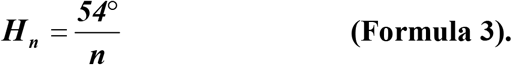

In this formula 54° is used as a constant value for each film width *n* in order to optimize the linear approximation of the *arctan* function (for details see error analysis in the supplementary supporting file).

Hence, Cobb’s angle *α* results as the product of the difference of the edge lengths *d* and the film’s characteristic parameter *H*_*n*_, which only depends on the film width *n*. In daily application, radiographs are usually taken with standardized film formats. Conveniently, factor *H*_*n*_ only needs to be calculated once for each film width *n* used. With *n = 15 cm, 20 cm and 35 cm* for example Formula 3 results in:

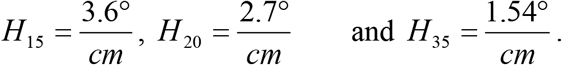

With fixed *H*_*n*_ only *d* needs to be determined for the calculation of *α* by Formula 2 (without a protractor).

For illustration refer to Figure 1:

Here from a (true to scale) print you can take *n = 7 cm* and 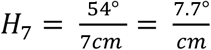.

For the determination of *α* by the Distance Method proceed as follows:

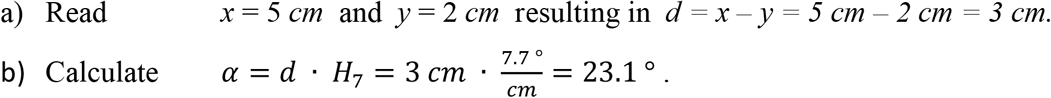

## 3. Comparison of the Three Methods

### 3.1 Error Analysis

The three methods considered here have in common that two tangent lines need to be established manually. Thus the variation of error in the determination of the tangent lines is identical for all methods. It is from the subsequent steps within each of the three different methods that different variations of error result for the whole procedures. Here the methods differ significantly in their variation of error and accuracy. In the supplementary file a detailed error analysis is given for the considered methods. As a result from this analysis we can state that with a probability of 95% the expected maximum deviation from the true angle occurring in steps 3 to 5 of the methods is limited as follows:

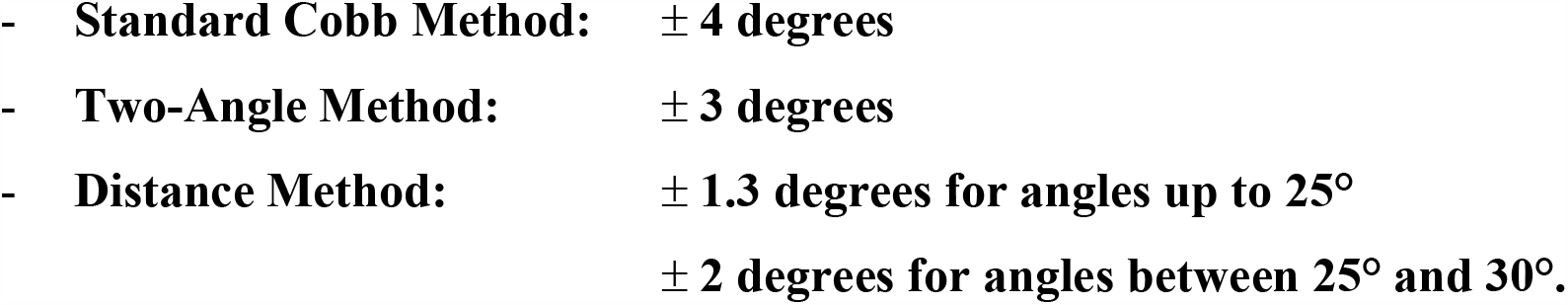

### 3.2 Test Measurements

In order to verify the different error deviations determined for the three methods by mathematical error analysis, test measurements were carried out with templates for different Cobb’s angles (in all templates a width of *n* = 18 cm has been used). For this purpose 60 test persons (students) were given 6 different measuring templates each, in which the end vertebrae lines for different geometric constellations were given (Cobb’s angles in the magnitudes of 5°, 10°, 20°, 30°, 40° and 50°). They were instructed to determine Cobb’s angle in every template pattern by the traditional Cobb Method (Four-Line Method) and to read the angles *β* and *γ* for the Two-Angle Method. Additionally, the line lengths *x* and *y* for the Distance Method should be read for all angles up to 30°. With the help of the formulas given for the different methods, the corresponding Cobb’s angles were then calculated on the basis of these values.

For all calculated angles, the mean values of the measurements from all three methods differed by less than 0.5° (as expected).

As a measure of the distribution of the data, the standard deviation of the 60 test results for Cobb’s angle was computed in each case for all procedures and every calculated angle. In the mean the three methods yielded the following standard deviations σ of the data:

In normally distributed samples 99.7% of the values are within a range of ± 3 *σ* of the mean value. Therefore we can conclude from the data that the variation of error of the mean value resulting from errors of measurement in steps 3 to 5 has the following magnitude:

For the recommended film width of *n* = 35 cm the standard deviation for the Distance Method (as presented in Table 1 for *n* = 18 cm) would halve proportionally to only 0.15 degrees, whereas it would remain unaltered for the two other methods. For this the 99.7% variation of error in the Distance Method even comes to only ± 0.45 degrees.

**Table 1:**
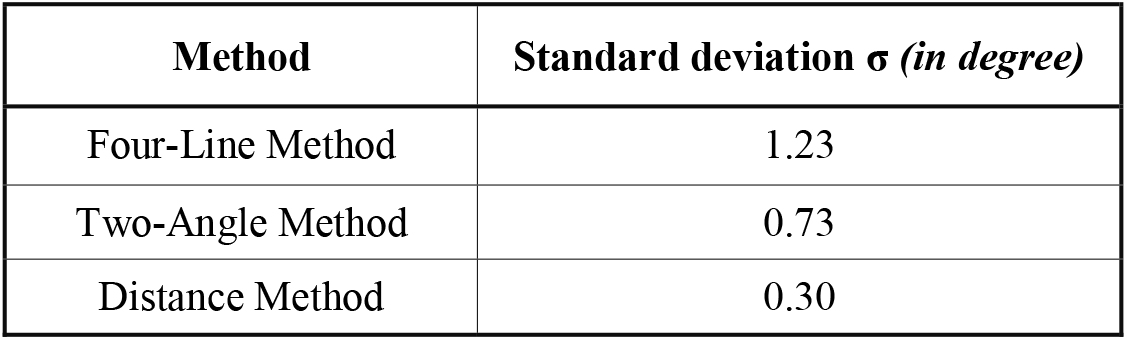
Standard Deviations of test measurements for the different calculation methods.

| Method | Standard deviation $\sigma$ (in degree) |
| --- | --- |
| Four-Line Method | 1.23 |
| Two-Angle Method | 0.73 |
| Distance Method | 0.30 |

**Table 2:**
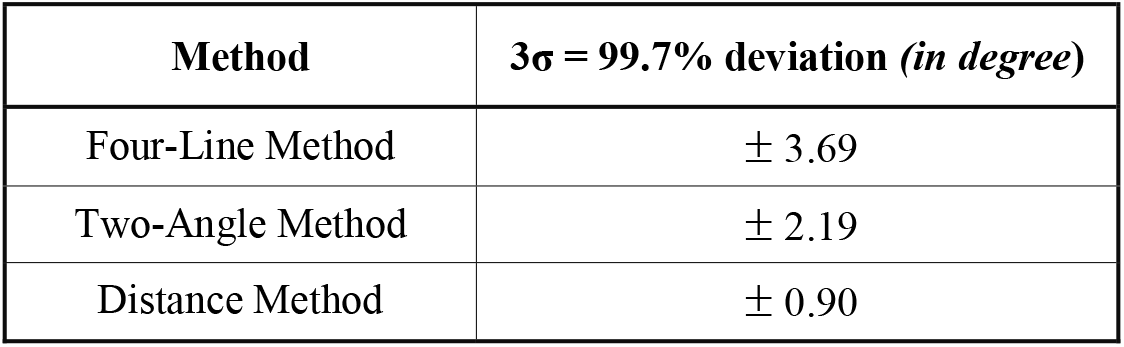
Error variations of test measurements for the different calculation methods.

| Method | $3\sigma = 99.7\%$ deviation (in degree) |
| --- | --- |
| Four-Line Method | $\pm 3.69$ |
| Two-Angle Method | $\pm 2.19$ |
| Distance Method | $\pm 0.90$ |

In their magnitude, the results of the test measurements confirm the proportion for the statistical distribution of the values obtained for the examined methods from error analysis.

### 3.3 Results

Compared to the Standard Cobb Method (Four-Line Method), the Two-Angle Method improves the variation of error for the subsequent steps 3 to 5 by at least 25 %. The Distance Method provides an improvement on the variation of error of at least 50 %, as long as Cobb’s angle does not exceed 30 degrees. In this method, the improvement on the variation of error is the better the wider the film format is. Using the Distance Method with a film width of 35 cm would even result in an improvement of 75 % for angles smaller than 25°. The advantage of accuracy of the Distance Method is especially evident for small Cobb’s angles as here the approximation error is almost zero and only the much smaller variation of the errors of measurement has an effect. When different Cobb angle measurements shall be compared for a patient over a time period for determination of changes in spine curvature, application of the distance method results in an additional gain of exactness. In this case the angles to be compared are of similar magnitude and the corresponding approximation errors from linearization of the *arctan* function are almost identical. Consequently, when subtracting these angles, the approximation error will be eliminated and the exactness of the difference will be influenced by the very small variation of measurement errors only.

Generally it can be stated, that for small angles the Distance Method has the least variation of error. Even if the approximation error resulting from linearization of the *arctan* function is included, it generates the best accuracy for angles up to 30 degrees. For larger angles, the Two-Angle Method provides a smaller but still significant improvement in the accuracy of the results compared to the traditional Cobb Method (Four-Line Method).

## 4. Suggestions for Practical Application

Whenever digitized imaging and calculation methods are not available and Cobb’s angle needs to be determined by hand on a radiograph, we recommend using the new procedures in order to minimize the variation of error. Both methods are more accurate, simpler, faster and more reliable than the traditional Four-Line Method.

For Cobb’s angles up to a magnitude of approx. 30 degrees, especially for small angles in the range between 1 and 10 degrees (e.g. in static scoliosis), the Distance Method should be preferred and for Cobb’s angles of more than 30 degrees the Two-Angle Method should be used instead of the Four-Line Method. The new methods can be applied also for the determination of any other curvature of the spine (lordosis, kyphosis).

For determination of Cobb’s angle in primary diagnostic of the lumbar spine (LS) we recommend using a radiograph format of 43 cm × 35 cm in which the LS and the pelvis can be pictured in a single image (in an upright position ap). Compared to an additional radiograph of the pelvis, the topographic advantages of this format are that all parameters of geometry and static of the pelvis and the LS may be determined more easily, more reliably, more quickly, more graphically and more accurately at a reduced x-ray exposure and lower costs. In particular, should the need arise, the level of scoliosis may be better correlated with a leg length difference (measured as a difference in height of the femur heads). However, the new methods of measuring and calculating are principally applicable for any film format.

## Data Availability

All relevant data are included in the manuscript.

